# Angiotensin I-Converting Enzyme type 2 expression is increased in pancreatic islets of type 2 diabetic donors

**DOI:** 10.1101/2023.06.25.23291752

**Authors:** Daniela Fignani, Erika Pedace, Giada Licata, Giuseppina Emanuela Grieco, Elena Aiello, Carmela de Luca, Lorella Marselli, Piero Marchetti, Guido Sebastiani, Francesco Dotta

## Abstract

**Aims:** Angiotensin I-converting enzyme type 2 (ACE2), a pivotal SARS-CoV-2 receptor, has been shown to be expressed in multiple cells including human pancreatic beta-cells. A putative bidirectional relationship between SARS-CoV-2 infection and diabetes has been suggested, confirming the hypothesis that viral infection in beta-cells may lead to new-onset diabetes or to a worse glycometabolic control in diabetic patients. However, whether ACE2 expression levels are altered in beta-cells of diabetic patients has not yet been investigated. Here, we aimed at elucidating the in-situ expression pattern of ACE2 in T2D respect to non-diabetic donors which may account for a higher susceptibility to SARS-CoV-2 infection in beta-cells.

**Material and methods:** ACE2 Immunofluorescence analysis using two antibodies alongside with insulin staining was performed on FFPE pancreatic sections obtained from n=20 T2D and n=20 non-diabetic multiorgan donors. Intensity and colocalization analyses were performed on a total of 1082 pancreatic islets. Macrophages detection was performed using anti-CD68 immunohistochemistry on serial sections from the same donors.

**Results:** Using two different antibodies, ACE2 expression was confirmed in beta-cells and in pancreas microvasculature. ACE2 expression was increased in pancreatic islets of T2D donors in comparison to non-diabetic controls alongside with a higher colocalization rate between ACE2 and insulin using both anti-ACE2 antibodies. CD68^+^ cells tend to be increased in T2D pancreata, in line with higher ACE2 expression observed in serial sections.

**Conclusions:** Higher ACE2 expression in T2D islets might increase their susceptibility to SARS-CoV-2 infection during COVID-19 in T2D patients, thus worsening glycometabolic outcomes and disease severity.

## Introduction

Previous studies have shown that patients with COVID-19 and pre-existing diabetes are at increased risk of being admitted to Intensive Care Unit (ICU), more likely to have multi-organ damage and had a higher mortality rate than non diabetic patients (1–4). In addition, abnormalities in glycaemic control, insulin resistance and pancreatic islet function have been observed in patients with COVID-19 without pre-existing history of diabetes, thus increasing the risk of developing Type 1 Diabetes (T1D), Type 2 Diabetes (T2D) or dysglycaemia (5–15). Therefore, it is conceivable to hypothesize a bidirectional relationship between COVID- 19 and diabetes (16,17).

Angiotensin I-Converting Enzyme type 2 (ACE2) is the canonical receptor for SARS-CoV-2. Entry of SARS-CoV-2 into cells depends on binding of the spike protein to ACE2. Hence, ACE2 expression levels dictates SARS-CoV-2 infectivity in multiple tissues [i.e. airway respiratory tract (18)], and is indispensable for SARS-CoV2 infection (19). Additional cofactors, such as TMPRSS2 and Neuropilin-1 (NRP-1) can facilitate and potentiate SARS-CoV-2 infection even in the presence of low levels of ACE2 (20).

The presence of SARS-CoV-2 receptor Angiotensin I-Converting Enzyme type 2 (ACE2) in pancreatic beta-cells was initially highly debated, with observed discrepancies among studies (21–23). At present, ACE2 expression in beta-cells is supported by multiple reports which clearly showed the expression of ACE2 and other co-factors (i.e. TMPRSS2, NRP1) in beta-cells (23–28). As a consequence, pancreatic islets are susceptible to SARS-CoV-2 infection mainly due to the expression of ACE2 and its co-factors, as demonstrated in several studies showing that beta-cells can be infected in vitro and in vivo in patients who died as a consequence of COVID-19 (24,25,27,28).

ACE2 expression has been shown to be modulated by pro-inflammatory stress with multiple studies showing increased expression of ACE2 upon exposure to several cytokines and/or pro-inflammatory molecules (23,29,30,31,32). Notably, chronic inflammation is a well-described feature of obesity and T2D (33–35). Inflammatory processes are also activated in pancreatic islets, as demonstrated by multiple evidence from animal models and humans with obesity and/or T2D (reviewed in 36). In this context, pancreatic islets low-grade inflammation has been shown to be associated with progressive beta-cell failure (37–39). In T2D, ACE2 expression has been shown to be increased in several organs exposed to a diabetic milieu (40–42). However, a systematic assessment of ACE2 expression in human pancreatic islets of T2D patients is still missing.

We here took advantage of the availability of a cohort of T2D and of non-diabetic multiorgan donors to investigate pancreatic islet ACE2 expression levels employing confocal immunofluorescence analysis and multiple antibodies directed against different ACE2 epitopes.

## Material and Methods

### Ethics Statement and multiorgan donors pancreata

Studies involving human participants were reviewed and approved by local ethics committee at the University of Pisa (Pisa, Italy). Pancreata not suitable for organ transplantation were obtained with informed written consent by organ donors’ next-of-kin and processed following standardized procedures. Human pancreatic tissue sections were obtained from pancreata of brain-dead adult non-diabetic and T2D multiorgan donors, COVID-19 negative. Multiple formalin-fixed paraffin embedded (FFPE) tissue sections were obtained from n = 20 non diabetic controls (CTR) [9F, 11M; age: 70.6 ± 7.0 years (mean ± S.D.); BMI: 26.2 ± 4.1 Kg/m^2^) and from n = 20 T2D pancreata (6F, 14M; age: 71.7±7.6 years (mean ± S.D.); BMI:27.1 ± 2.7 Kg/m^2^ (mean ± S.D.)] whose clinical characteristics are described in **ESM Table 1**.

**Table 1.** Main clinical characteristics of ND and T2D subjects included in the study.

| CLINICAL PARAMETERS | ND<br>(n = 20) | T2D<br>(n = 20) | *p* value |
| --- | --- | --- | --- |
| Age | 70,6±7.05 | 71.6±7.7 | ns |
| Sex (M:F) | 11:9 | 14:6 | ns |
| BMI (kg/m<sup>2</sup>) | 26.2±4.2 | 27.1±2.7 | ns |
| Abdominal circumference (cm) | 103.0±20.7 | 100±10.3 | ns |
| ICU stay (days) | 3.9±5.0 | 3.4±1.9 | ns |
| Cold Ischemia Time (hours) | 16.0±5.29 | 15.76±9.29 | ns |
| Mean glycaemia (mmol/L) | 8.03±2.0 | 11.6±3.7 | *p* < 0.05 |
| Diabetes Duration (years) | - | 10.9±8.8 | n/a |

### Immunofluorescence and Immunohistochemical analysis of human pancreatic sections

#### Triple immunofluorescence for ACE2 (MAB933)-ACE2 (AB15348)-Insulin

Formalin-Fixed Paraffin Embedded (FFPE) tissue sections (7 μm thick) were obtained from pancreata of non-diabetic or T2D multiorgan donors. FFPE sections were analyzed using quadruple immunofluorescence for INS, ACE2 (two different antibodies) and DAPI for nuclei counterstain.

After deparaffinization and rehydration (xylene-I and xylene II, 20min/each; ethanol 100%, 95%, 80%, 75%, all vol./vol., 5min/each), pancreatic sections were subjected to antigen retrieval through incubation in 10 mM citrate buffer [0.1 M Citric Acid Monohydrate (Cat. C1909 - Sigma Aldrich, St. Louis, MO, USA) and 0.1 M Tribasic Sodium Citrate Dihydrate (cat. S4641 -Sigma Aldrich, St. Louis, MO, USA)] pH 6.0 in microwave (600 Watt) for 10 minutes, maintaining boiling conditions. After cooling and 3 washes in PBS 1X (Cat. 14040- 091 - Gibco, ThermoFisherScientific, Waltham, MA, USA), sections were incubated with PBS 1X supplemented with 3% Bovine Serum Albumin (BSA) (cat. A1470-25G - Sigma Aldrich, St. Louis, MO, USA) for 40 minutes at RT to avoid non-specific reactions.

Then, sections were incubated with primary antibody monoclonal mouse anti-human ACE2 (cat. MAB933 - R&;D System, Minneapolis, MS, USA) (final concentration: 15 μg/ml) in PBS 1X supplemented with 3% BSA and primary antibody polyclonal rabbit anti-human ACE2 (cat. ab15348 - Abcam, Cambridge, UK) (final concentration: 0,5 μg/ml) in PBS 1X supplemented with 3% BSA, overnight at +4°C. After 3 washes in PBS 1X, the sections were incubated for 1h at RT with ready to use polyclonal guinea pig anti-human insulin (cat. IR002-Agilent Technologies, Santa Clara, CA, USA) further diluted 1:5 in PBS 1X supplemented with 3% BSA. Then, the sections were incubated 1 hour with related secondary antibodies, all diluted 1:500 (final concentration: 4 μg/ml) in PBS 1X: goat anti-guinea pig Alexa-Fluor 555 conjugate (cat. A21435, Molecular Probes, ThermoFisher Scientific, Waltham, MA, USA); goat anti-mouse Alexa-Fluor 488 conjugate (cat. A11029, Molecular Probes, ThermoFisher Scientific, Waltham, MA, USA) and goat anti-rabbit 647 conjugate (cat. A21245 – Molecular Probes, ThermoFisher Scientific, Waltham, MA, USA). Sections were counterstained with 4′,6-Diamidino-2-phenylindoledihydrochloride (DAPI, cat. D8517, Sigma-Aldrich) diluted 1:3000 in PBS 1X and then mounted with Dako Fluorescence Mounting Medium (cat. S3023 – Agilent Technologies, Santa Clara, CA, USA). The sections were stored overnight at +4°C until image analysis.

To confirm the validity of the staining 1 μg of polyclonal rabbit anti-human ACE2 (cat. ab15348 -Abcam, Cambridge, UK) was combined with or without 10 μg of the immunizing human ACE2 peptide (cat. 15325 – Abcam, Cambridge, UK) and the staining was performed as describe above.

#### Double immunofluorecence for ACE2 (MAB933)-CD31

After deparaffinization and rehydration, FFPE pancreatic sections were subjected to antigen retrieval, as described above.

After cooling and 3 washes in PBS 1X (Cat. 14040-091 - Gibco, ThermoFisher Scientific, Waltham, MA, USA), sections were incubated with PBS 1X supplemented with 3% Bovine Serum Albumin (BSA) (cat. A1470-25G - Sigma Aldrich, St. Louis, MO, USA) for 40 minutes at RT to avoid non-specific reactions and then incubated overnight at +4°C with primary antibody monoclonal rabbit anti-human CD31 (cat. Ab76533 - Abcam, Cambridge, UK) (final concentration: 3,5 µg/ml). After 3 washes in PBS 1X, the sections were incubated with monoclonal mouse anti-human ACE2 (cat. MAB933 - R&D System, Minneapolis, MS, USA) (final concentration: 15 µg/ml) for 1 hour at RT.

After 3 washes in PBS 1X, the sections were incubated for 1 hour at RT with ready to use polyclonal guinea pig anti-human insulin (cat. IR002 - Agilent Technologies, Santa Clara, CA, USA) further diluted 1:5 in PBS 1X supplemented with 3% BSA. Then, sections were incubated 1 hour with related secondary antibodies, all diluted 1:500 (final concentration: 4 μg/ml) in PBS 1X: goat anti-guinea pig Alexa-Fluor 594 conjugate (cat. A11037, Molecular Probes, ThermoFisher Scientific, Waltham, MA, USA); goat anti-mouse Alexa-Fluor 488 conjugate (cat. A11029, Molecular Probes, ThermoFisher Scientific, Waltham, MA, USA) and goat anti-rabbit 647 conjugate (cat. A21245 –Molecular Probes, ThermoFisher Scientific, Waltham, MA, USA). Sections were counterstained with 4′,6-Diamidino-2- phenylindole dihydrochloride (DAPI, cat. D8517, Sigma Aldrich, St. Louis, MO, USA) diluted 1:3000 in PBS 1X and then mounted with Dako Fluorescence Mounting Medium (cat. S3023 – Agilent Technologies, Santa Clara, CA, USA). The sections were stored overnight at +4°C until image analysis.

#### CD68-positive cells immunohistochemical staining

Pancreatic CD68-positive cells were detected using enzymatic-colorimetric immunohistochemical staining. After deparaffinization and rehydration (see above), pancreatic sections were subjected to blocking of peroxidase with 3% hydrogen peroxide in PBS 1X for 20 minutes. Then, the sections were subjected to heat-induced antigen retrieval using Tris-EDTA buffer (10 mmol/l Tris, 1 mmol/l EDTA, 0.05% Tween-20, pH 9.0) for 20min at 100°C. After cooling and incubation in 3% BSA in PBS 1X for 30min at RT to reduce non-specific reactions, sections were stained in 3% BSA in PBS 1X for 1h at RT with mouse monoclonal anti-human CD68 (cat. M0876 - Agilent Technologies, Santa Clara, CA, USA) (final concentration: 0,4 mg/ml). After three washes in PBS 1X, sections were incubated with secondary antibody polyclonal goat anti-mouse HRP-conjugate (cat.115-036-003 - Jackson ImmunoResearch, Philadelphia, PA, USA) diluted 1:500 in PBS 1X for 1h at RT. Subsequently, the sections were incubated with one drop of 3,3-Diaminobenzidine (DAB) chromogen solution (cat. RE7270-K, Novolink MAX DAB, Leica Microsystems, Wetzlar, Germany) for 5 minutes, to trigger the colorimetric reaction. After 10 minutes of incubation in water, the sections were incubated for 1 hour at RT with ready to use polyclonal guinea pig anti-human insulin (cat. IR002 - Agilent Technologies, Santa Clara, CA, USA) further diluted 1:5 in PBS 1X supplemented with 3% BSA. After three washes in PBS 1X, sections were incubated with secondary polyclonal antibody goat anti-Guinea Pig conjugated with Alkaline Phosphatase (AP) (cat. A18772- ThermoFisher Scientific, Waltham, MA, USA) (final concentration: 0,3 µg/ml). Subsequently, the sections were incubated with one drop of Liquid Fast Red (cat. K0640 – Agilent Technologies, Santa Clara, CA, USA) (a drop of chromogen in 3 ml of substrate Levamisole (cat.X3021 - Agilent Technologies, Santa Clara, CA, USA) one drop per ml of LFR of Levamisole) for 5 minutes.

Stained sections were then counterstained with hematoxylin (cat. MHS31 - Sigma Aldrich, St. Louis, MO, USA) for 4 minutes. After 1 hour of air dry the sections were covered with a drop of Faramount, Aqueous Mounting Medium, Ready-to-Use (cat. S302580-2 - Agilent Technologies, Santa Clara, CA, USA).

### Image analysis

Images were acquired, as a single stack focal plane, employing a Leica TCS SP5 confocal laser scanning microscope system (Leica Microsystems, Wetzlar, Germany).

For confocal laser scanning microscope system sections were scanned and images acquired at 40× magnification. The same confocal microscope setting parameters were applied to all stained sections before image acquisition, in order to uniformly collect detected signal related to each channel.

Colocalization analysis between ACE2 and insulin was performed using LasAF software (Leica Microsystems, Wetzlar, Germany). The region of interest (ROI) was drawn to calculate the colocalization rate (which indicates the extent of colocalization between two different channels and reported as a percentage) as a ratio between the colocalization area and the image foreground. Evaluation of ACE2 expression intensity in human pancreatic islets was performed using LasAf software (www.leica-microsystem.com). This software calculates the ratio between intensity sum ROI (which indicates the sum ROI of the grey-scale value of pixels within a region of interest) of ACE2 channel and Area ROI (μm^2^) of human pancreatic islets. Both in colocalization and intensity measurement analysis, a specific threshold was assigned based on the fluorescence background. The same threshold was maintained for all the images in all the cases analyzed.

Insulin positive area was measured using Volocity 6.3 software (Perkin Elmer, Waltham, MA, USA). Relative insulin signal positive area was calculated as a ratio between Area Intensity sum ROI (μm^2^) of insulin channel and Area ROI (μm^2^) of each human pancreatic islets.

For CD68-positive cells detection and quantification, images of the entire section were acquired using NanoZoomer S60 Digital slide scanner (cat. C13210-01 - Hamamatsu Photonics, Hamamatsu City, Japan) and were displayed using the proprietary NDP.view2 software. Manual count of CD68^+^ cells on the entire section area was performed.

### Statistical analysis

Results were expressed as mean ± Standard Deviation (S.D.). Comparisons between two groups were carried out using Mann-Whitney U test for non-parametric data (normality checked using Kolgomorov-Smirnov test). Differences were considered significant with p values less than 0.05. Clinical variable associations with ACE2 expression were checked using multiple least square regression analysis. Statistical analyses were performed using Graph Pad Prism 8 software.

## Results

To detect pancreatic ACE2 protein expression and distribution, and to evaluate differences between non diabetic (ND) and type 2 diabetic (T2D) donors, we performed a quadruple immunofluorescence analysis on FFPE pancreatic sections obtained from n = 20 ND and n = 20 T2D multiorgan donors (**Table 1 and ESM Table 1**). To cross-validate the ACE2 staining results, we used two different anti-ACE2 antibodies: (*i*) a monoclonal mouse IgG2a anti-human ACE2 (R&D, MAB933), whose specificity was confirmed through an isotype primary antibody staining (**ESM Figure 1A**) and (*ii*) a rabbit polyclonal anti-ACE2 (Abcam, Ab15348), whose specificity was tested through a peptide competition assay and subsequent staining in ND donors pancreatic sections (**ESM Figure 1A**).

**Figure 1.**
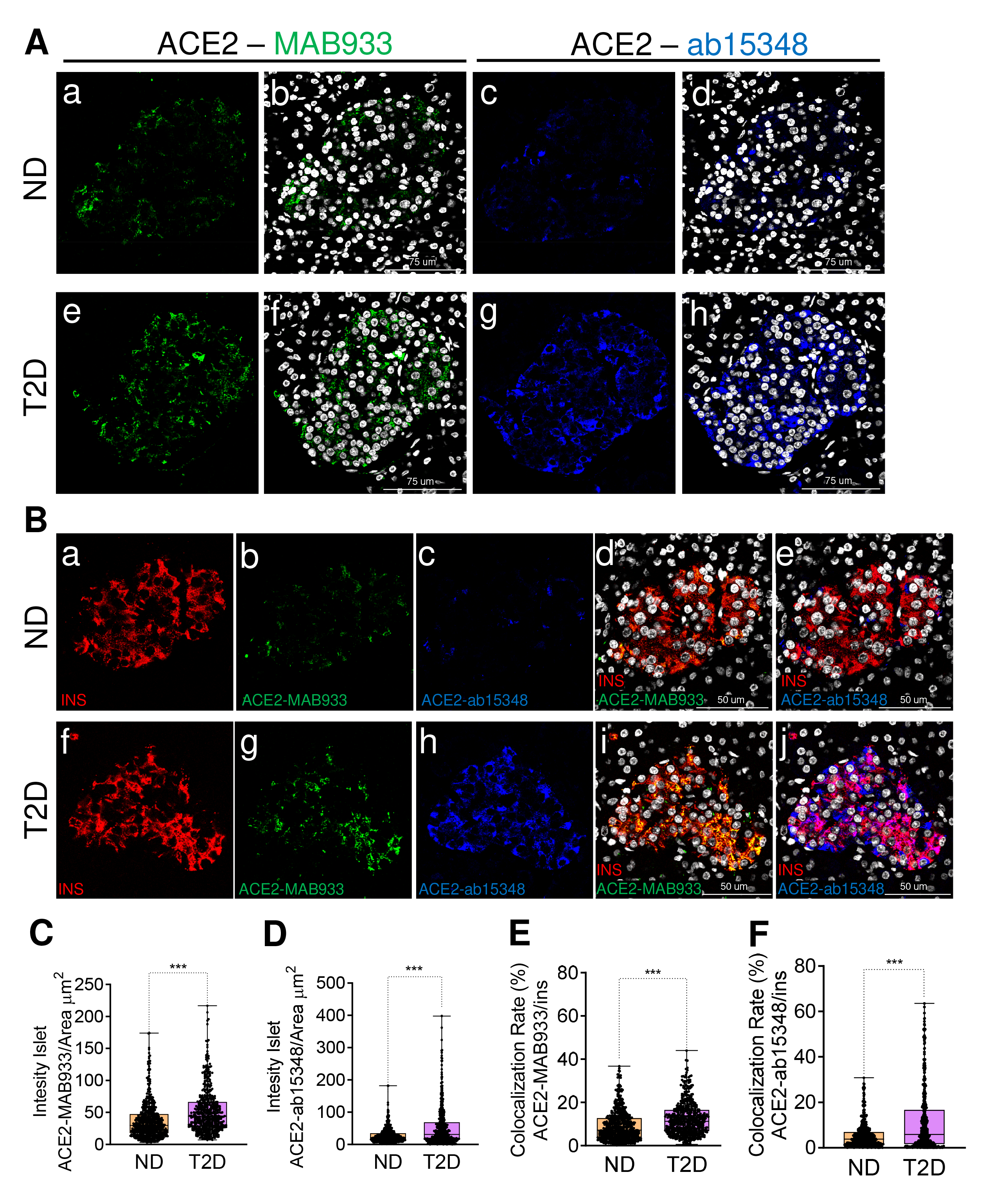
Upregulation of ACE2 in pancreatic islet beta-cells of T2D donors. (**A**) Representative confocal images of FFPE pancreatic section from nondiabetic (ND) (case 13) and T2D donor (case 38). Pancreatic sections were stained with anti-ACE2 Ab MAB933 (green, panels a and e) and the merge with DAPI (white, nuclei) (panels b and f) showing the expression of ACE2-MAB933 in a pancreatic islet; pancreatic sections were stained with anti-ACE2 Ab ab15348 (blue, panels c and g) and the merge with DAPI (white, nuclei) (panel d and h) showed the expression of ACE2-ab15348 in a pancreatic islet. (**B**) Representative confocal images of FFPE pancreatic section derived from a nondiabetic (ND) (case 2) and a T2D donor (case 32). Pancreatic sections were stained for insulin (INS, red, panels a and f), ACE2-MAB933 (green, panels b and g) and ACE2-ab15348 (blue, panels c and h). Colocalization between ACE2-MAB933 and insulin is showed in yellow (panels d and i), while colocalization between ACE2-ab15348 and insulin is reported in magenta (panels e and j). Signal intensity analysis measured with anti-ACE2-MAB933 (**C**) and with anti-ACE2- ab15348 (**D**) antibody in non diabetic and T2D pancreatic sections; values are shown as fluorescence intensity of each islet detected (ND=556 islets; T2D=526 islets) reported as the sum of gray-scale values for each pixel normalized for the islets area (ROI, mm^2^). **(E-F)** Colocalization rate analysis between ACE2-MAB933-Insulin (**E**) and ACE2-ab15348-insulin (**F**). Values are shown the colocalization rate (ROI, mm^2^). \**p* < 0.05, \*\**p* < 0.01, \*\*\**p* < 0.001, non-parametric Mann-Whitney U test, performed after checking normality with the Kolmogorov-Smirnov normality test.

In ND and T2D donor pancreata, both antibodies showed signals indicating that ACE2 is expressed in pancreatic islets where it is mostly colocalized with insulin signal (**ESM Figure 2A**). A triple staining on ND FFPE pancreatic sections using ACE2-MAB933, insulin and glucagon antibodies confirmed the prevalent colocalization of ACE2 with insulin in comparison to ACE2 with glucagon (**ESM Figure 2A**), in line with previous data (23). The present results confirm that ACE2, in pancreatic islets, is prevalent in beta-cells as expected.

**Figure 2.**
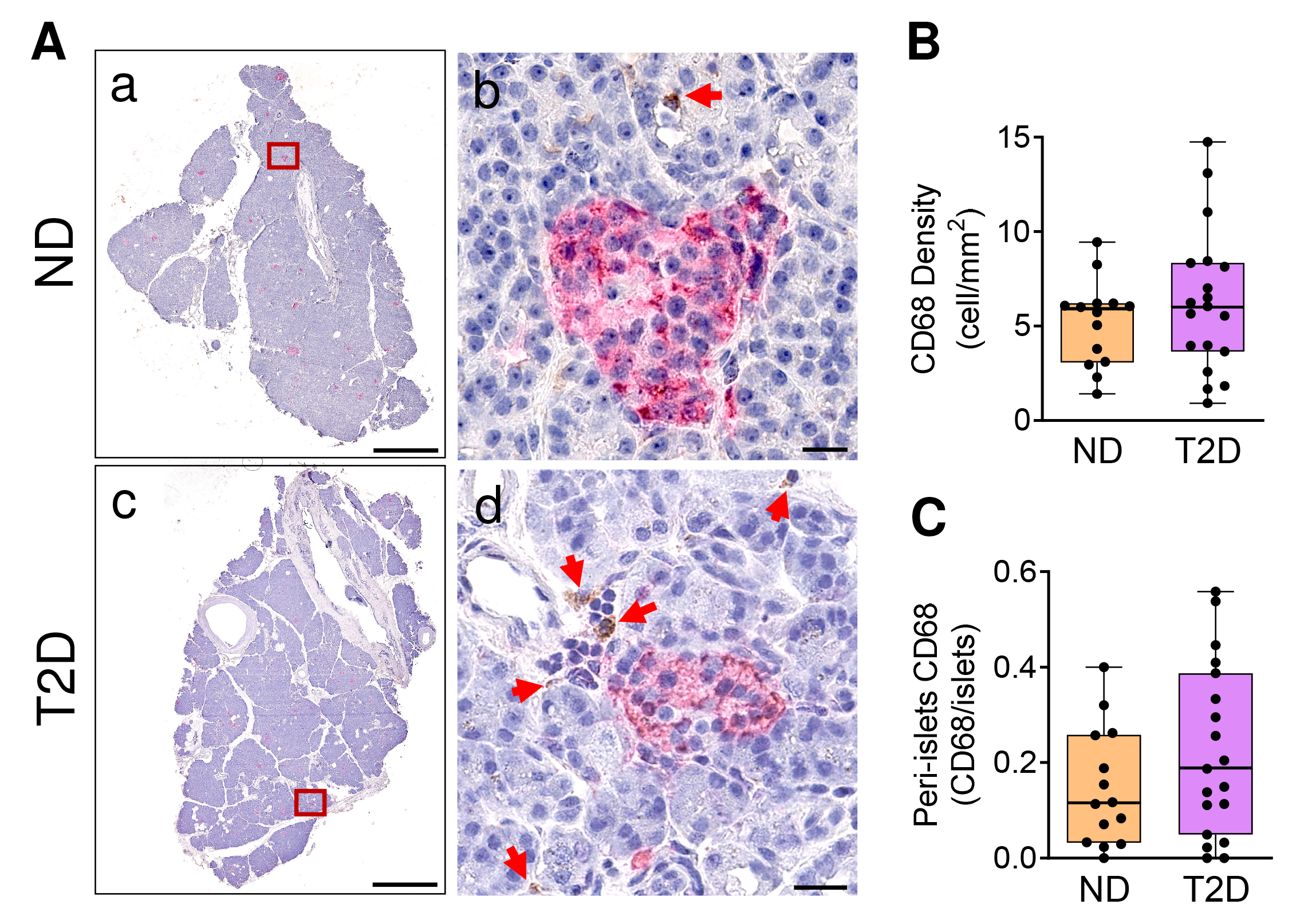
Peri-islets CD68^+^-macrophages in T2D pancreas. **A)** Representative whole-slide image of FFPE pancreatic section from ND (panel a) and T2D donor pancreatic tissue sections (panel c) stained for insulin (red) and CD68 (brown). Zoom-in insets of the peri-islets CD68+ macrophages are reported in panel b (ND) and in panel d (T2D). **B)** CD68 density in the whole pancreas is reported as cell/mm^2^. Peri-islets CD68+-macrophages (**C**), considered within 250 µm from islet edge, are reported as number of positive cells per islet.

Outside pancreatic islets, ACE2-positive cells showed a vasculature-like morphology and distribution; such results were confirmed by using two different ACE2 antibodies (**ESM Figure 3A**); a subsequent co-staining with ACE2 and vascular-endothelial marker CD31 showed the juxtaposition of the two signals (**ESM Figure 3A**), thus confirming our previous observations (23), in line with other reports (22), which demonstrated the localization of ACE2 also in pancreatic vascular cells.

Next, we focused on ACE2 expression in pancreatic islets. To evaluate putative ACE2 expression differences between ND and T2D, we performed an analysis of the intensity of ACE2 signals including a total of n=1082 islets. Both antibodies revealed a higher intensity of ACE2 in T2D compared to ND pancreatic islets (**Figure 1A**). Analysis of ACE2 intensity confirmed the significantly increased expression of ACE2 in T2D pancreatic islets compared to ND donors as measured by R&D and Abcam antibodies (greyscale values of ACE2- MAB933 in T2D=52.5±34.6 and in ND =37.1±28.1, *p* < 0.001; greyscale values of ACE2- ab15348 in T2D=53.2±63.5 and in ND =27.3±22.3, *p* < 0.001) (**Figure 1C, 1D**).

Since ACE2 expression in pancreatic islets is mostly prevalent in beta-cells, we performed a colocalization rate analysis (reported as the percentage of the overlap of INS and ACE2 signals) of ACE2 MAB933/INS and ACE2 Ab15348/INS in T2D and ND pancreatic islets. We observed a significantly increased colocalization rate between ACE2 and INS in T2D compared to ND subjects (**Figure 1B**). The ACE2 colocalization rate analysis of n=1082 islets across all ND and T2D donors confirmed the increased colocalization rate between ACE2-MAB933/INS [T2D: 12.5 ± 7.6% (mean ± S.D.) vs ND 9.23 ± 7.0% (mean ± S.D.), *p* < 0.001] and of ACE2-ab15348/INS [T2D: 11.45 ± 13.7% (mean±S.D.) vs ND 5.3% ± 5.3%; *p* < 0.001] in T2D compared to ND (**Figure 1E and Figure 1F**). No differences in insulin-positive area and signals were observed between T2D and ND donors (**ESM Figure 4**). In the multiple linear regression analysis (**ESM Table 2**), ACE2 expression (reported as staining intensity) was not associated with age, BMI, gender, ICU stay or duration of cold ischaemia time, thus excluding the influence of these putative confounding variables on ACE2 levels. Of note, ACE2 expression was not associated with gender or blood glucose levels (**ESM Table 2**).

To investigate a possible link between ACE2 expression and inflammation triggered by innate immune cells in the pancreas of T2D donors, we analysed CD68^+^ macrophages in pancreatic tissue. In the whole pancreatic section (**Figure 2A**), CD68^+^ macrophages showed an abundance of about 6.2 cells per mm^2^ in T2D and 5.1 cells per mm^2^ in ND. Interestingly, CD68^+^ macrophages in the peri-islets showed an increased abundance trend in T2D compared to ND donors (0.22 vs. 0.15 CD68+ cells/islet) (**Figure 2B**); although not statistically significant, this result is consistent with the increased expression of ACE2 observed in pancreatic islets of previous serial sections.

Overall, these data show an increased expression of ACE2 in pancreatic beta cells in T2D compared to ND donors. Although such increase is independent of available clinical variables related to glycometabolic outcomes, we observed a tendency to increase in peri-islets CD68^+^-macrophages thus putatively associating ACE2 expression increase to inflammatory insults.

## Discussion

The bidirectional relationship between SARS-CoV-2 infection and diabetes mellitus has been hypothesised since the beginning of the pandemic. As a matter of fact, evidence of hyperglycaemia and abnormalities in glycometabolic control after SARS-CoV-2 infection were demonstrated in several studies (5–15); moreover, diabetic patients showed a more severe outcome, a poor prognosis and higher mortality rate after SARS-CoV-2 infection (1,2,5–8). Notably, it has been demonstrated that SARS-CoV-2 can infect human host cells through the binding of viral spike protein to the extracellular N-terminal domain of the ACE2 receptor (24,25). As a matter of fact, we and others have previously shown that beta cells do express ACE2, thus being susceptible to SARS-CoV-2 infection (23–28).

In this study, we analysed an extended cohort of T2D multiorgan donors in comparison to age-and sex-matched non diabetic ones, to evaluate ACE2 expression and distribution. We demonstrated that ACE2 is increased in pancreatic islets of T2D donors and showed a higher colocalization rate in beta cells of T2D versus ND donors. Notably, we considered a total of n=1082 pancreatic islets across all ND and T2D donors, and the results were obtained using two different anti-ACE2 antibodies (monoclonal ACE2-MAB933 from R&D and polyclonal ACE2 Ab1538 from Abcam) adopted in the immunofluorescence analysis in an experimental cross-validation approach. The higher colocalization rate between ACE2 and insulin in T2D suggests that, in pancreatic islets, ACE2 hyperexpression is mainly occurring in beta-cells; however, at this stage, we cannot decipher whether (*i*) ACE2 expression is increased in beta-cells already expressing the receptor, (*ii*) it is increased due to de-novo expression occurring in ACE2-negative beta-cells, or (*iii*) a combination of both mechanisms. Additional analyses using imaging machine learning approaches and single cells segmentation are required to further decipher the intra-islet expression pattern of ACE2 in T2D. Overall, our data support an increased expression of the SARS-CoV2 receptor in beta-cells of T2D donors, and are in-line with a recent report demonstrating the upregulation of ACE2 in pancreatic islets of T2D donors subjected to microarray and RNA sequencing (43); Indeed, Taneera and colleagues showed that ACE2 is elevated in diabetic islets but no correlation between its expression and HbA1c, age or BMI was detected, similarly to what we have observed in the present study. In contrast to our results, other previous reports did not observe the upregulation of ACE2 in T2D islets (21,22); this can be due to the high heterogeneity of ACE2 expression or differences among T2D cohorts and/or reagents adopted. It is worth noting that in the present study we adopted two different antibodies after a detailed analysis of their specificity and efficiency testing.

Previous studies showed that other organs exposed to a diabetic milieu such as lung, kidney and heart showed the upregulation of ACE2, thus corroborating our findings in a different context (40–42). Indeed, ACE2 expression was found increased in bronchial epithelium and alveolar tissue of T2D donors and a linear relationship was detected between blood glucose levels and ACE2 expression in alveolar tissue (40). In the heart tissue, ACE2 expression was significantly increased in cardiomyocytes of T2D patients with poor glycaemic control respect to ND patients and T2D patients with good glycaemic control (41). In kidney organoids ACE2 was expressed in tubular-like cells and an oscillatory glucose regimen induced the expression of ACE2 (42). Collectively, we can hypothesise that ACE2 expression is increased upon exposure to inflammation and/or high glucose or other stressors and that such chronic stress stimuli also exert their deleterious effect on beta cells favouring the upregulation of ACE2. However, unlike other reports, we cannot find a significant correlation between ACE2 expression and blood glucose levels. This can be explained by the high level of glycaemia already observed in ND patients during the ICU stay (Table 1); alternatively, we can hypothesize that, at least in beta-cells, high glucose is not the main factor leading to the hyperexpression of ACE2 and that pro-inflammatory molecules may play a major role in ACE2 modulation. The latter hypothesis is also supported by Van der Heide and colleagues who did not find any association between ACE2 expression and high glucose exposure in beta-cells (26). In addition, our previous study showed that the in vitro exposure of the beta-cell line EndoC-βH1 or primary pancreatic islets to pro-inflammatory molecules (i.e. IFNγ+IL-1β+TNFα or IFN α), but not metabolic stressors such as palmitate, can significantly increase ACE2 expression (23), thus supporting the hypothesis of a major inflammatory-mediated mechanism governing ACE2- hyperexpression in beta-cells.

In line with this hypothesis, we explored the potential contribution of inflammation in ACE2 upregulation in T2D pancreatic islets, by analysing pancreatic-tissue resident CD68^+^- macrophages. We observed an increase of peri-islets CD68^+^ macrophages, even though not significant. However, such increase is supported by other previous observations which showed a peri-islets increase of CD68^+^-macrophages in T2D pancreata in comparison to ND donors (44,45). Thus, additional analyses should be considered to further explore the inflammatory mechanisms leading to ACE2 upregulation in beta cells in T2D.

It has been suggested that increased expression of ACE2 may explain the increased infectivity or severity of COVID-19 in patients with diabetes (46). In this context, we can argue that increased expression of ACE2 in beta cells may lead to increased susceptibility of beta-cells to SARS-CoV-2 infection, making beta cells more prone to virus tropism in patients already infected with SARS-CoV-2. However, further analyses are needed to explore the expression of ACE2 cofactors (i.e. NRP1, TMPRSS2) in T2D islets and to decipher their contribution in the putative enhancement of the susceptibility of beta-cells to SARS-CoV-2 infection during COVID-19.

In conclusion, we observed the upregulation of ACE2 in pancreatic islet beta-cells of T2D donors, putatively driven by inflammatory-mediated mechanisms. Higher ACE2 expression in T2D islets might increase their susceptibility to SARS-CoV-2 infection during COVID-19 disease in T2D patients, thus exacerbating glycometabolic outcomes and worsening the severity of the disease.

## Data Availability

All data produced in the present study are available upon reasonable request to the authors

## Acknowledgements

The secretarial help of Maddalena Prencipe and Alessandra Mechini is greatly appreciated. We thank Dr. Noemi Brusco, for her expertise and support in the image analysis.

## Data availability

The data generated in this study are available from the corresponding authors upon request.

## Funding

The work is supported by the Innovative Medicines Initiative 2 (IMI2) Joint Undertaking under grant agreement No.115797-INNODIA and No.945268 INNODIA HARVEST. This joint undertaking receives support from the Union’s Horizon 2020 research and innovation programme and EFPIA, JDRF and the Leona M. and Harry B. Helmsley Charitable Trust. FD was supported by the Italian Ministry of University and Research (2017KAM2R5_003). GS was supported by the Italian Ministry of University and Research (201793XZ5A_006).

## Authors’ relationships and activities

The authors declare that the research was conducted in the absence of any commercial or financial relationships that could be construed as a potential conflict of interest.

## Contribution statement

All authors contributed substantially to the conception and design, data acquisition and analysis, interpretation and drafting the article or revising it critically. All authors approved the final version of the manuscript and its content. DF, GS and FD are guarantors of this work and as such, had full access to all the data in the study and take responsibility for the integrity of the data and the accuracy of data analyses.

## Notes

### Competing Interest Statement

The authors have declared no competing interest.

### Author Declarations

Local ethics committee of University of Pisa (Pisa, Italy) gave ethical approval for this work

## References

1. Myers LC, Parodi SM, Escobar GJ, Liu VX. Characteristics of Hospitalized Adults With COVID-19 in an Integrated Health Care System in California. JAMA 2020;323:2195–2198.

2. Barron E, Bakhai C, Kar P, Weaver A, Bradley D, Ismail H, et al. Associations of type 1 and type 2 diabetes with COVID-19-related mortality in England: a whole-population study. Lancet Diabetes Endocrinol 2020;8:813–822.

3. Chen N, Zhou M, Dong X, Qu J, Gong F, Han Y, et al. Epidemiological and clinical characteristics of 99 cases of 2019 novel coronavirus pneumonia in Wuhan, China: a descriptive study. Lancet 2020;395:507–513.

4. Fadini GP, Morieri ML, Longato E, Avogaro A. Prevalence and impact of diabetes among people infected with SARS-CoV-2. J Endocrinol Invest 2020;43:867–869.

5. Wander PL, Lowy E, Beste LA, Tulloch-Palomino L, Korpak A, Peterson AC, et al. The Incidence of Diabetes Among 2,777,768 Veterans With and Without Recent SARS-CoV-2 Infection. Diabetes Care 2022;45:782–788.

6. Birabaharan M, Kaelber DC, Pettus JH, Smith DM. Risk of new-onset type 2 diabetes in 600 055 people after COVID-19: A cohort study. Diabetes Obes Metab 2022;24:1176–1179.

7. Guo Y, Bian J, Chen A, Wang F, Posgai AL, Schatz DA, et al. Incidence Trends of New-Onset Diabetes in Children and Adolescents Before and During the COVID-19 Pandemic: Findings from Florida. Diabetes 2022.

8. Sardu C, D’Onofrio N, Balestrieri ML, Barbieri M, Rizzo MR, Messina V, et al. Outcomes in Patients With Hyperglycemia Affected by COVID-19: Can We Do More on Glycemic Control? Diabetes Care 2020;43:1408–1415.

9. Kendall EK, Olaker VR, Kaelber DC, Xu R, Davis PB. Association of SARS-CoV-2 Infection With New-Onset Type 1 Diabetes Among Pediatric Patients From 2020 to 2021. JAMA Netw Open 2022;5:e2233014.

10. McKeigue PM, McGurnaghan S, Blackbourn L, Bath LE, McAllister DA, Caparrotta TM, et al. Relation of Incident Type 1 Diabetes to Recent COVID-19 Infection: Cohort Study Using e-Health Record Linkage in Scotland. Diabetes Care 2022.

11. Kamrath C, Rosenbauer J, Eckert AJ, Siedler K, Bartelt H, Klose D, et al. Incidence of Type 1 Diabetes in Children and Adolescents During the COVID-19 Pandemic in Germany: Results From the DPV Registry. Diabetes Care 2022;45:1762–1771.

12. Xie Y, Al-Aly Z. Risks and burdens of incident diabetes in long COVID: a cohort study. Lancet Diabetes Endocrinol 2022;10:311–321.

13. Cromer SJ, Colling C, Schatoff D, Leary M, Stamou MI, Selen DJ, et al. Newly diagnosed diabetes vs. pre-existing diabetes upon admission for COVID-19: Associated factors, short-term outcomes, and long-term glycemic phenotypes. J Diabetes Complicat 2022;36:108145.

14. Shulman R, Cohen E, Stukel TA, Diong C, Guttmann A. Examination of Trends in Diabetes Incidence Among Children During the COVID-19 Pandemic in Ontario, Canada, From March 2020 to September 2021. JAMA Netw Open 2022;5:e2223394.

15. Montefusco L, Nasr M Ben, D’Addio F, Loretelli C, Rossi A, Pastore I, et al. Acute and long-term disruption of glycometabolic control after SARS-CoV-2 infection. Nat Metab 2021;3:774–785.

16. Groß R, Kleger A. COVID-19 and diabetes - where are we now? Nat Metab 2022.

17. Khunti K, Prato S Del, Mathieu C, Kahn SE, Gabbay RA, Buse JB. COVID-19, Hyperglycemia, and New-Onset Diabetes. Diabetes Care 2021;44:2645–2655.

18. Hou YJ, Okuda K, Edwards CE, Martinez DR, Asakura T, Dinnon KH, et al. SARS-CoV-2 Reverse Genetics Reveals a Variable Infection Gradient in the Respiratory Tract. Cell 2020;182:429–446.e14.

19. Hoffmann M, Kleine-Weber H, Schroeder S, Krüger N, Herrler T, Erichsen S, et al. SARS-CoV-2 Cell Entry Depends on ACE2 and TMPRSS2 and Is Blocked by a Clinically Proven Protease Inhibitor. Cell 2020;181:271–280.e8.

20. Cantuti-Castelvetri L, Ojha R, Pedro LD, Djannatian M, Franz J, Kuivanen S, et al. Neuropilin-1 facilitates SARS-CoV-2 cell entry and infectivity. Science 2020;370:856– 860.

21. Kusmartseva I, Wu W, Syed F, Der Heide V Van, Jorgensen M, Joseph P, et al. Expression of SARS-CoV-2 Entry Factors in the Pancreas of Normal Organ Donors and Individuals with COVID-19. Cell Metab 2020;32:1041–1051.e6.

22. Coate KC, Cha J, Shrestha S, Wang W, Gonçalves LM, Almaça J, et al. SARS-CoV-2 Cell Entry Factors ACE2 and TMPRSS2 Are Expressed in the Microvasculature and Ducts of Human Pancreas but Are Not Enriched in β Cells. Cell Metab 2020;32:1028– 1040.e4.

23. Fignani D, Licata G, Brusco N, Nigi L, Grieco GE, Marselli L, et al. SARS-CoV-2 Receptor Angiotensin I-Converting Enzyme Type 2 (ACE2) Is Expressed in Human Pancreatic β-Cells and in the Human Pancreas Microvasculature. Front Endocrinol (Lausanne) 2020;11:596898.

24. Müller JA, Groß R, Conzelmann C, Krüger J, Merle U, Steinhart J, et al. SARS-CoV-2 infects and replicates in cells of the human endocrine and exocrine pancreas. Nat Metab 2021;3:149–165.

25. Steenblock C, Richter S, Berger I, Barovic M, Schmid J, Schubert U, et al. Viral infiltration of pancreatic islets in patients with COVID-19. Nat Commun 2021;12:3534.

26. van der Heide V, Jangra S, Cohen P, Rathnasinghe R, Aslam S, Aydillo T, et al. Limited extent and consequences of pancreatic SARS-CoV-2 infection. Cell Rep 2022;38:110508.

27. Wu C-T, Lidsky PV, Xiao Y, Lee IT, Cheng R, Nakayama T, et al. SARS-CoV-2 infects human pancreatic β cells and elicits β cell impairment. Cell Metab 2021;33:1565– 1576.e5.

28. Qadir MMF, Bhondeley M, Beatty W, Gaupp DD, Doyle-Meyers LA, Fischer T, et al. SARS-CoV-2 infection of the pancreas promotes thrombofibrosis and is associated with new-onset diabetes. JCI Insight 2021;6.

29. Ziegler CGK, Allon SJ, Nyquist SK, Mbano IM, Miao VN, Tzouanas CN, et al. SARS-CoV-2 Receptor ACE2 Is an Interferon-Stimulated Gene in Human Airway Epithelial Cells and Is Detected in Specific Cell Subsets across Tissues. Cell 2020;181:1016– 1035.e19.

30. Kimura H, Francisco D, Conway M, Martinez FD, Vercelli D, Polverino F, et al. Type 2 inflammation modulates ACE2 and TMPRSS2 in airway epithelial cells. J Allergy Clin Immunol 2020;146:80–88.e8.

31. Sajuthi SP, DeFord P, Li Y, Jackson ND, Montgomery MT, Everman JL, et al. Type 2 and interferon inflammation regulate SARS-CoV-2 entry factor expression in the airway epithelium. Nat Commun 2020;11:5139.

32. Busnadiego I, Fernbach S, Pohl MO, Karakus U, Huber M, Trkola A, et al. Antiviral Activity of Type I, II, and III Interferons Counterbalances ACE2 Inducibility and Restricts SARS-CoV-2. MBio 2020;11.

33. Donath MY, Shoelson SE. Type 2 diabetes as an inflammatory disease. Nat Rev Immunol 2011;11:98–107.

34. DeFronzo RA, Ferrannini E, Groop L, Henry RR, Herman WH, Holst JJ, et al. Type 2 diabetes mellitus. Nat Rev Dis Primers 2015;1:15019.

35. Joya-Galeana J, Fernandez M, Cervera A, Reyna S, Ghosh S, Triplitt C, et al. Effects of insulin and oral anti-diabetic agents on glucose metabolism, vascular dysfunction and skeletal muscle inflammation in type 2 diabetic subjects. Diabetes Metab Res Rev 2011;27:373–382.

36. Böni-Schnetzler M, Meier DT. Islet inflammation in type 2 diabetes. Semin Immunopathol 2019;41:501–513.

37. Eguchi K, Manabe I, Oishi-Tanaka Y, Ohsugi M, Kono N, Ogata F, et al. Saturated fatty acid and TLR signaling link β cell dysfunction and islet inflammation. Cell Metab 2012;15:518–533.

38. Ying W, Lee YS, Dong Y, Seidman JS, Yang M, Isaac R, et al. Expansion of Islet-Resident Macrophages Leads to Inflammation Affecting β Cell Proliferation and Function in Obesity. Cell Metab 2019;29:457–474.e5.

39. Maedler K, Dharmadhikari G, Schumann DM, Størling J. Interleukin-1 beta targeted therapy for type 2 diabetes. Expert Opin Biol Ther 2009;9:1177–1188.

40. Wijnant SRA, Jacobs M, Eeckhoutte HP Van, Lapauw B, Joos GF, Bracke KR, et al. Expression of ACE2, the SARS-CoV-2 Receptor, in Lung Tissue of Patients With Type 2 Diabetes. Diabetes 2020;69:2691–2699.

41. Marfella R, D’Onofrio N, Mansueto G, Grimaldi V, Trotta MC, Sardu C, et al. Glycated ACE2 reduces anti-remodeling effects of renin-angiotensin system inhibition in human diabetic hearts. Cardiovasc Diabetol 2022;21:146.

42. Garreta E, Prado P, Stanifer ML, Monteil V, Marco A, Ullate-Agote A, et al. A diabetic milieu increases ACE2 expression and cellular susceptibility to SARS-CoV-2 infections in human kidney organoids and patient cells. Cell Metab 2022;34:857–873.e9.

43. Taneera J, El-Huneidi W, Hamad M, Mohammed AK, Elaraby E, Hachim MY. Expression Profile of SARS-CoV-2 Host Receptors in Human Pancreatic Islets Revealed Upregulation of ACE2 in Diabetic Donors. Biology (Basel) 2020;9.

44. Wu M, Lee MYY, Bahl V, Traum D, Schug J, Kusmartseva I, et al. Single-cell analysis of the human pancreas in type 2 diabetes using multi-spectral imaging mass cytometry. Cell Rep 2021;37:109919.

45. Ehses JA, Perren A, Eppler E, Ribaux P, Pospisilik JA, Maor-Cahn R, et al. Increased number of islet-associated macrophages in type 2 diabetes. Diabetes 2007;56:2356– 2370.

46. Roberts J, Pritchard AL, Treweeke AT, Rossi AG, Brace N, Cahill P, et al. Why Is COVID-19 More Severe in Patients With Diabetes? The Role of Angiotensin-Converting Enzyme 2, Endothelial Dysfunction and the Immunoinflammatory System. Front Cardiovasc Med 2020;7:629933.

