## Supplementary Material for "Angiotensin I-Converting Enzyme type 2 expression is increased in pancreatic islets of type 2 diabetic donors"

### ELECTRONIC SUPPLEMENTARY MATERIAL (ESM)

**ESM Table 1.** Available main clinical characteristics for each non-diabetic (ND) and type 2 diabetic (T2D) donors included in the study. Gender, Age range (years), BMI (kg/m$^2$), Abdominal circumference (cm) and disease duration (years) are reported.

| Pancreas ID | Gender (M/F) | Age range (y) | BMI (Kg/m2) | Abdominal circumference (cm) | Years from diagnosis of diabetes | ICU stay (days) | Cold Ischemia Time (hours) | Mean glycemia (mg/dl) |
|---|---|---|---|---|---|---|---|---|
| 1 | F | 61-65 | 23,3 | 90 | / | 1 | 15 | 152 |
| 2 | F | 61-65 | 23,53 | 87 | / | 3 | 15 | 163 |
| 3 | M | 61-65 | 27,8 | 116 | / | 12 | 14 | 252 |
| 4 | F | 66-70 | 20,8 | 80 | / | 3 | 13 | 138 |
| 5 | M | 71-75 | 24,83 | 100 | / | 2 | 13 | 161 |
| 6 | M | 76-80 | 23,15 | 94 | / | 1 | 14 | 115 |
| 7 | M | 71-75 | 34,6 | 130 | / | 9 | 12 | 130 |
| 8 | F | 76-80 | 19,53 | 90 | / | 2 | 17 | 66 |
| 9 | M | 81-85 | 27,68 | 107 | / | 2 | 18 | 110 |
| 10 | F | 71-75 | 34,89 | 145 | / | 5 | 16 | 181 |
| 11 | F | 56-60 | 27,68 | 126 | / | 2 | 17 | 134 |
| 12 | F | 61-65 | 29,38 | 115 | / | 2 | 20 | 159 |
| 13 | F | 61-65 | 23,4 | 108 | / | 1 | n/a | 159 |
| 14 | M | 71-75 | 32,98 | 120 | / | 1 | 36 | 146 |
| 15 | M | 66-70 | 25 | 87 | / | 6 | 17 | 148 |
| 16 | M | 76-80 | 27,8 | 132 | / | 21 | 13 | 160 |
| 17 | M | 66-70 | 25,05 | 85 | / | 1 | 12 | 111 |
| 18 | M | 81-85 | 25,7 | 89 | / | 1 | 13 | 127 |
| 19 | F | 71-75 | 23,44 | 63 | / | 1 | 16 | 155 |
| 20 | M | 61-65 | 24,69 | 110 | / | 1 | 14 | 126 |
| 21 | F | 71-75 | 22,2 | 90 | 9 | 1 | 11 | 155 |
| 22 | M | 76-80 | 27,3 | 105 | 5 | 2 | 15 | 200 |
| 23 | M | 66-70 | 25,9 | 88 | 11 | 2 | 15 | 334 |
| 24 | M | 76-80 | 25,9 | 103 | 25 | 2 | 13 | 203 |
| 25 | M | 66-70 | 27,8 | 105 | 6 | 9 | 11 | 234 |
| 26 | F | 51-55 | 33,1 | 117 | 7 | 2 | 12 | 138 |
| 27 | F | 71-75 | 25,39 | 90 | n/a | 2 | n/a | 130 |
| 28 | M | 76-80 | 27,18 | 110 | n/a | 2 | n/a | 210 |
| 29 | F | 76-80 | 29,4 | 105 | 2 | 1 | 13 | 153 |
| 30 | M | 61-65 | 33,1 | 118 | 5 | n/a | 12 | 171 |
| 31 | M | 76-80 | 26,1 | 97 | 30 | 3 | 12 | n/a |
| 32 | M | 56-60 | 29,4 | 80 | 4 | 1 | 12 | 181 |
| 33 | M | 71-75 | 26,1 | 110 | n/a | 1 | 15 | 374 |
| 34 | M | 61-65 | 24,6 | 92 | 10 | 2 | 14 | 177 |
| 35 | F | 76-80 | 29,2 | 110 | 6 | n/a | 29 | 269 |
| 36 | M | 76-80 | 26 | 95 | 14 | 2 | n/a | 165 |
| 37 | M | 76-80 | 24,5 | 90 | 9 | 4 | 48 | 303 |
| 38 | F | 71-75 | 27,3 | 108 | 30 | 1 | 11 | 214 |
| 39 | M | 66-70 | 27,7 | 97 | 5 | 5 | 13 | 151 |
| 40 | M | 76-80 | 24,7 | 106 | 8 | 1 | 12 | 211 |

**ESM Table 2.** Multiple least square regression analysis to identify putative correlations between ACE2 expression in pancreatic islets and clinical characteristics available in ND and T2D donors. "Age range (years)", "BMI (Kg/m$^2$)", "Abdominal circumference (cm)", "ICU stay (days)", "Cold Ischemia Time (hours)", "Mean glycemia (mg/dl)" and "Gender (M/F)[M]" were included as independent variable while ACE2 expression measured using MAB933 (**a**) and Ab15348 (**b**) as dependent variables in two separate models. *Estimate* values report the increase (positive values) or decrease (negative values) of ACE2 expression for each unit of the independent variable.

**a) Multiple least square linear regression analysis predicting dependency to pancreatic islets ACE2 expression measured using Ab MAB933 antibody**

| Independent Variable | Estimate | Standard error | 95% CI (asymptotic) | P value |
|---|---|---|---|---|
| Age (years) | 0,09676 | 0,6393 | -1,220 to 1,414 | 0,8809 |
| Gender (M/F)[M] | 13,46 | 10,03 | -7,204 to 34,12 | 0,1918 |
| BMI (Kg/m2) | 1,824 | 1,831 | -1,947 to 5,596 | 0,3287 |
| Abdominal circumference (cm) | -0,4605 | 0,405 | -1,295 to 0,3736 | 0,2663 |
| ICU stay (days) | 0,05898 | 1,209 | -2,432 to 2,550 | 0,9615 |
| Cold Ischemia Time (hours) | -1,09 | 0,6228 | -2,373 to 0,1922 | 0,0922 |
| Mean glycemia (mg/dl) | -0,0366 | 0,07049 | -0,1818 to 0,1086 | 0,6083 |

**b) Multiple least square linear regression predicting dependency to pancreatic islets ACE2 expression measured using Ab15348 antibody**

| Independent Variable | Estimate | Standard error | 95% CI | P value |
|---|---|---|---|---|
| Age (years) | 1,112 | 1,184 | -1,326 to 3,550 | 0,3566 |
| Gender (M/F)[M] | -15,02 | 18,58 | -53,28 to 23,24 | 0,4265 |
| BMI (Kg/m2) | 2,871 | 3,391 | -4,113 to 9,855 | 0,4052 |
| Abdominal circumference (cm) | -0,9164 | 0,75 | -2,461 to 0,6283 | 0,2332 |
| ICU stay (days) | 1,441 | 2,239 | -3,172 to 6,053 | 0,5259 |
| Cold Ischemia Time (hours) | -1,067 | 1,153 | -3,443 to 1,308 | 0,3636 |
| Mean glycemia (mg/dl) | 0,09076 | 0,1305 | -0,1781 to 0,3596 | 0,4933 |

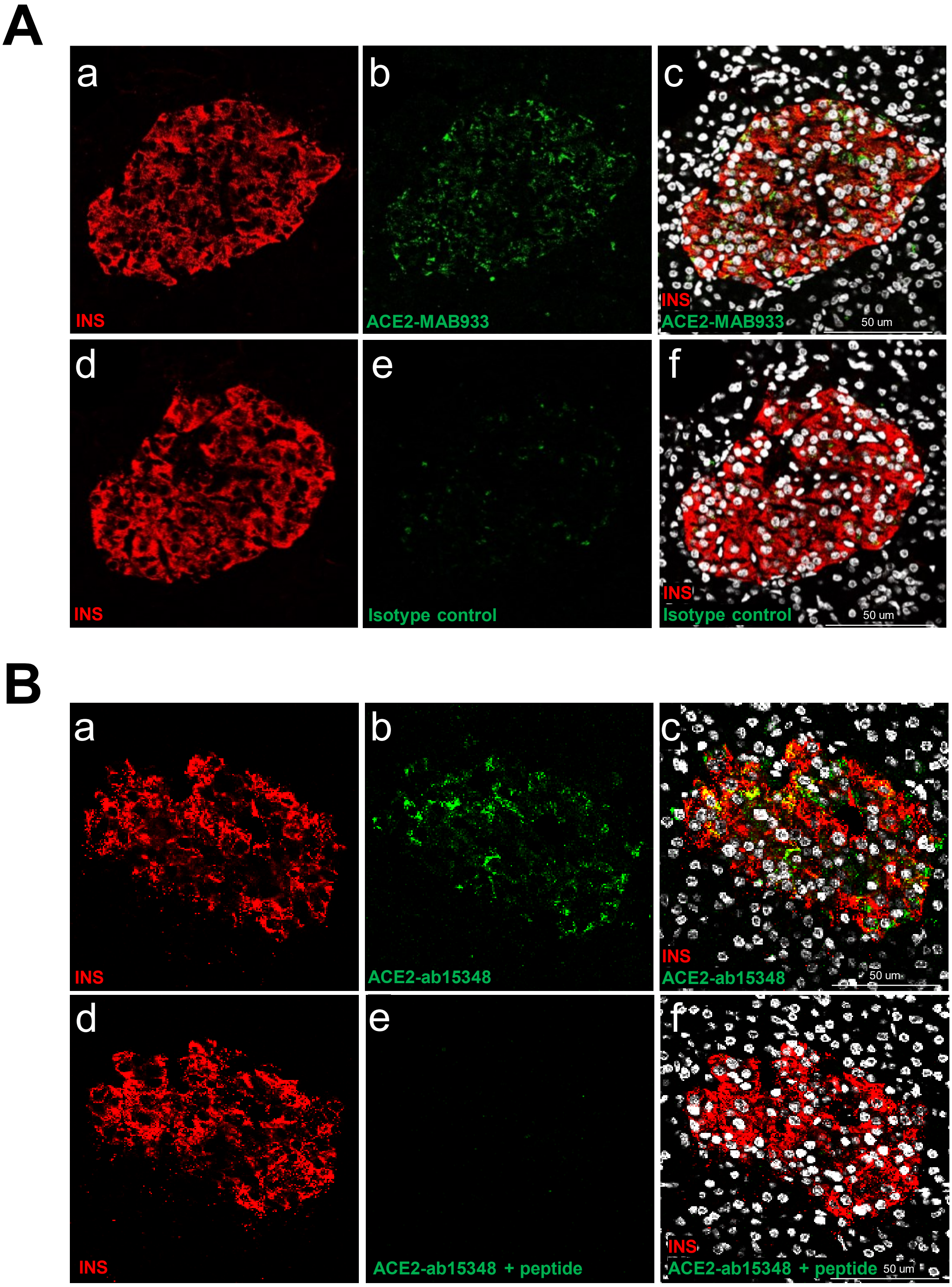


**ESM Figure 1**. **A) Isotype antibody negative control staining**. In panels a-c, representative confocal image of negative control of the immunofluorescence analysis of insulin/ACE2-MAB933 using an isotype primary antibody. A FFPE pancreatic section from a ND donor were stained for insulin (INS, red, panel a) and ACE2-MAB933 (green, panel b) and merged signals showing colocalization between insulin and ACE2-MAB933 in yellow (panel c). In panels d-f, representative confocal image of the serial FFPE pancreatic section showed in panels a-c, and stained for insulin (INS, red, panel d) and ACE2-MAB933 isotype antibody control (panel e). Merged cannels (panel

f) showing no signal for ACE2 and no colocalization between insulin and ACE2. Scale bars in panel c and f is 50 µm.

**B**) **Peptide competition assay negative control**. In panels a-c, representative confocal image of the immunofluorescence analysis of insulin/ACE2 on FFPE pancreatic sections stained for insulin (INS, red, panel a) and ACE2-ab15348 (green, panel c) and merged signals showing colocalization between insulin and ACE2-ab15348 in yellow (panel c). In panels d-f, representative confocal images of the serial FFPE pancreatic section showed in panel a-c, and stained for insulin (INS, red, panel D) and ACE2 ab15348 pre-incubated with an ACE2 peptide (panel e) showing no positive signal. Merged channels (panel f) showing no signal for ACE2 and no colocalization between insulin and ACE2. Scale bars in panel c and f is 50 µm.

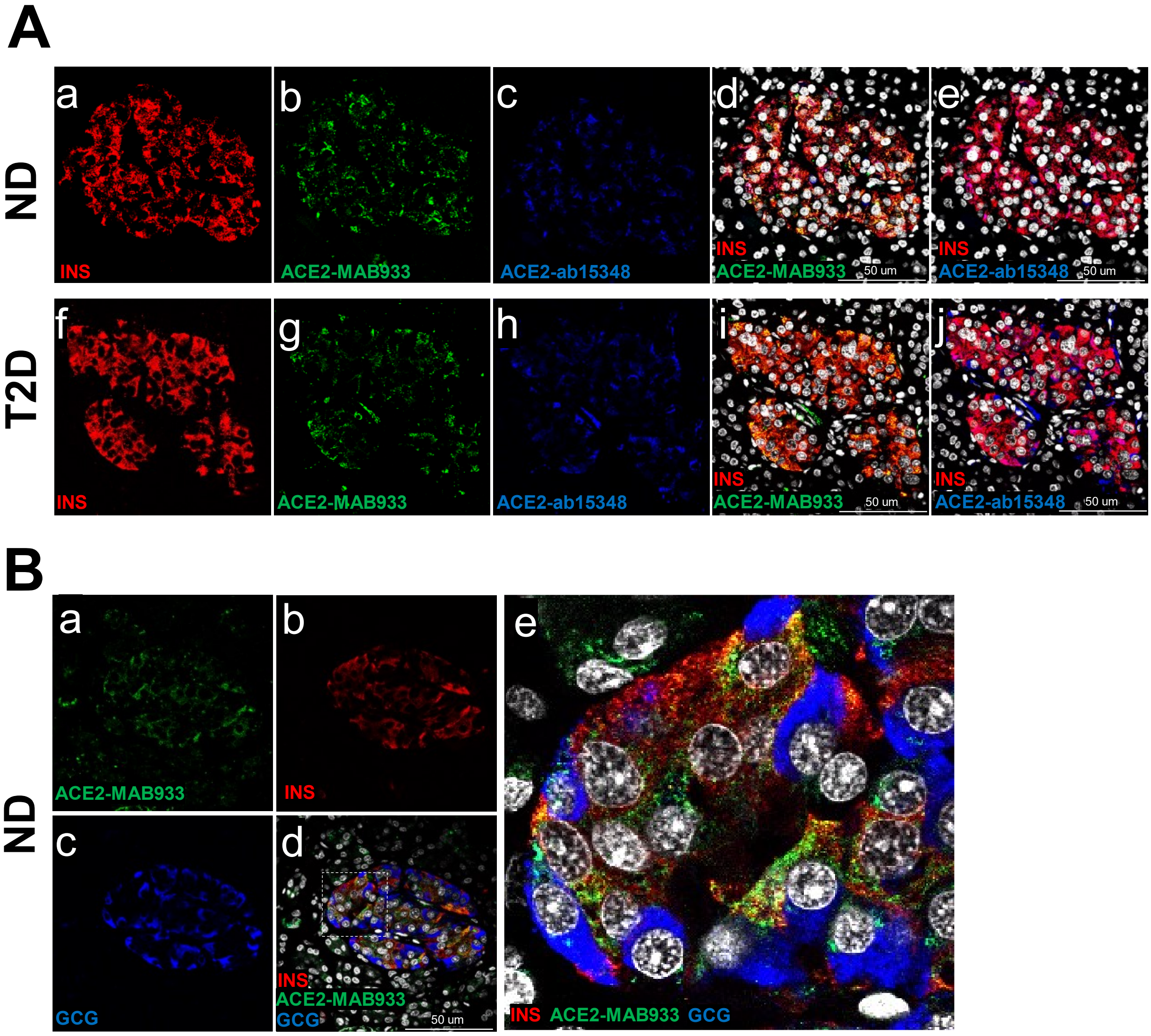


**ESM Figure 2. ACE2 expression pattern in human pancreatic islets. A)** Representative confocal images of ACE2-MAB933, ACE2-Ab15348 and insulin staining in pancreatic islet in FFPE pancreatic sections derived from a non diabetic case (ND) (panels a-e) and from a T2D case (panels f-j). Tissues were stained for DAPI (white), insulin (INS, red, panel a, f), ACE2-MAB933 (green, panels b, g) and ACE2-ab15348 (blue, panels c, h). Colocalization between insulin and ACE2-MAB933 is showed in yellow (panels d, i) and colocalization between insulin and ACE2-ab15348 is reported in magenta (panels e, j).

**B)** Confocal images of ACE2-MAB933, insulin and glucagon staining in a FFPE pancreatic section derived from a non-diabetic donor (ND). A FFPE pancreatic section was stained with DAPI (nuclei in white), ACE2-MAB933 (green, panel a), insulin (INS, red, panel b) and glucagon (GCG, blue, panel c). Merged images were shown in panel d and in Zoom-in inset in panel e. Scale bar in panels d is 50 μm.

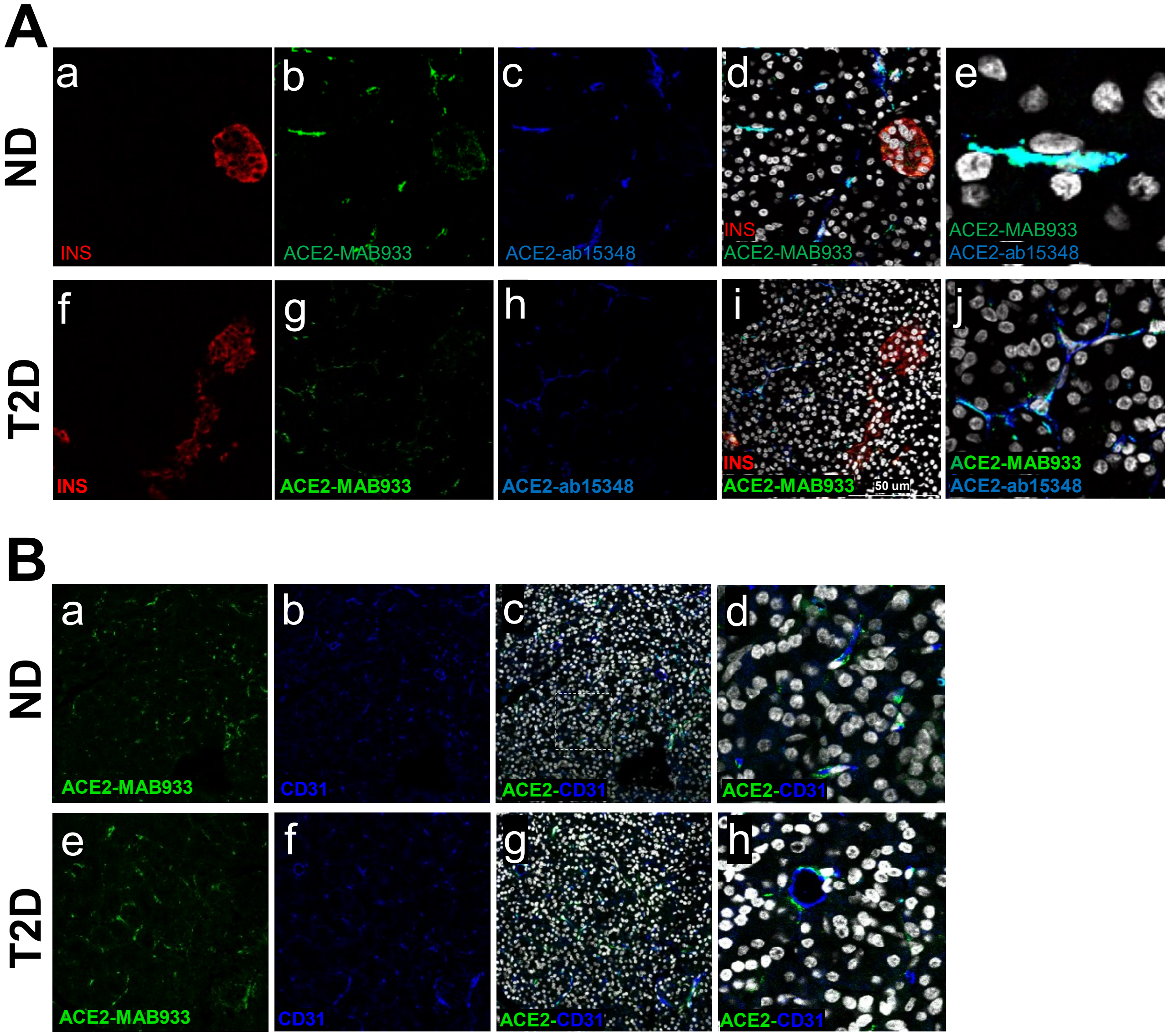


**ESM Figure 3. ACE2 expression pattern in human pancreas microvasculature.** Confocal images of ACE2 in pancreas microvasculature in FFPE pancreatic section derived from a non diabetic case (ND) and from a T2D case. In **A)**, tissues were stained for DAPI (white), insulin (INS, red, panel a, f), ACE2-MAB933 (green, panels b, g) and ACE2-ab15348 (blue, panels C, H). Colocalization between ACE2-MAB933 and ACE2-ab15348 in cells outside pancreatic islets showed a vasculature like-morphology (panel d, e, i, j). A Zoom-in inset (in panels e, j) shows details of ACE2 expression in microvasculature-like cells in ND (panel e) and in T2D (panel j). Scale bars in panels D and I are 50 µm.

In **B**), tissues were stained for DAPI (white), ACE2-MAB933 (green, panels a, e) and microvasculature endothelial marker CD31 (blue, panels b, f). Merged images are shown in panels c and g. Zoom-in insets are reported in panels d and h.

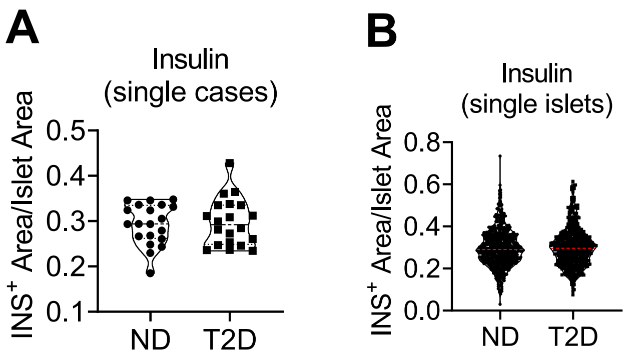


**ESM Figure 4. Insulin-positive area analysis on FFPE pancreatic sections of non-diabetic and type 2 diabetic multiorgan donors. A)** Imaging analysis of insulin-positive signal area normalized per islet area in n=20 non-diabetic (ND) and n=20 T2D donors. Each dot represents the mean insulin-positive area/islet area for all islets analysed in each case. **B**) Imaging analysis of insulin-positive signal area normalized per islet area, in n=556 islets from non-diabetic (ND) donors and n=529 islets from T2D donors. Data are shown as violin plot with median value dotted lines.
